# Group testing for SARS-CoV-2 allows for up to 10-fold efficiency increase across realistic scenarios and testing strategies

**DOI:** 10.1101/2020.04.30.20085290

**Authors:** Claudio M. Verdun, Tim Fuchs, Pavol Harar, Dennis Elbrächter, David S. Fischer, Julius Berner, Philipp Grohs, Fabian J. Theis, Felix Krahmer

## Abstract

**Background:** Due to the ongoing COVID-19 pandemic, demand for diagnostic testing has increased drastically, resulting in shortages of necessary materials to conduct the tests and overwhelming the capacity of testing laboratories. The supply scarcity and capacity limits affect test administration: priority must be given to hospitalized patients and symptomatic individuals, which can prevent the identification of asymptomatic and presymptomatic individuals and hence effective tracking and tracing policies. We describe optimized group testing strategies applicable to SARS-CoV-2 tests in scenarios tailored to the current COVID-19 pandemic and assess significant gains compared to individual testing.

**Methods:** We account for biochemically realistic scenarios in the context of dilution effects on SARS-CoV-2 samples and consider evidence on specificity and sensitivity of PCR-based tests for the novel coronavirus. Because of the current uncertainty and the temporal and spatial changes in the prevalence regime, we provide analysis for a number of realistic scenarios and propose fast and reliable strategies for massive testing procedures.

**Findings:** We find significant efficiency gaps between different group testing strategies in realistic scenarios for SARS-CoV-2 testing, highlighting the need for an informed decision of the pooling protocol depending on estimated prevalence, target specificity, and high-vs. low-risk population. For example, using one of the presented methods, all 1·47 million inhabitants of Munich, Germany, could be tested using only around 141 thousand tests if the infection rate is below 0·4% is assumed. Using 1 million tests, the 6·69 million inhabitants from the city of Rio de Janeiro, Brazil, could be tested as long as the infection rate does not exceed 1%.

**Interpretation:** Altogether this work may help provide a basis for efficient upscaling of current testing procedures, taking the population heterogeneity into account and fine grained towards the desired study populations, e.g. cross-sectional versus health-care workers and adapted mixtures thereof.

**Funding:** German Science Foundation (DFG), German Federal Ministry of Education and Research (BMBF), Chan Zuckerberg Initiative DAF and Austrian Science Fund (FWF).

**Research in context:** *Evidence before this study:* The concept of group testing goes back to mathematical ideas developed in the 1940’s and has already been successfully implemented for various infectious diseases but also in non-medical settings such as testing for failures of electronic components. The issue of group testing for SARS-CoV-2 has been addressed in a number of very recent papers and preprints including feasibility studies of different laboratories and a few methodological overview papers. Nevertheless, to the best of our knowledge, no study provided a comprehensive comparison contrasting hierarchical testing, array testing, and informative testing strategies based on combined groups for stratified populations and relying on up-to-date data about the accuracy of PCR-based test – all of them feasible to be implemented in the current pandemic. Moreover, a discussion of massive informative testing strategies for pandemic scenarios, employing combined pools consisting of high-risk and low-risk individuals, was still missing in the public health literature.

*Added value of this study:* We analyse sensitivity, specificity, and throughput of group testing schemes in a series of scenarios tailored to realistic prevalence rates for SARS-CoV-2 in stratified populations and to the characteristics of the qRT-PCR tests used to diagnose COVID-19. Our analysis yields a comprehensive characterisation of a wide range of pooling schemes, broken down by population characteristics, that can serve as a guideline to be queried by testing entities to meet their settings. In particular, our findings demonstrate that a promising strategy to test asymptomatic or presymptomatic individuals in conjunction with high-risk individuals such as healthcare workers is to amalgamate them together in a suitable way, and we provide adequate pool configurations. Such strategies had not been explored for SARS-CoV-2 testing as of yet. We also provide insights on group testing under constraints, i.e. when the number of stages, maximum group size and false negative rate of the whole method are restricted to a certain range of realistic values. We introduce efficient parallelizable non-adaptive test procedures for simplified and fast large-scale test design in case of severe shortage of test components. We develop an intuitive web application that can be used by any researcher working on the front line of testing procedures to visualize all the different strategies and to design pooling schemes in an flexible manner according to their specific prevalence scenario and test configuration.

*Implications of all the available evidence:* Testing for SARS-CoV-2 presents new challenges to authorities such as rapidly changing prevalence estimates and bottlenecks in testing capacity. In this context, applying an appropriately chosen group testing procedure can allow for up to 10-fold increase of the feasible throughput, such that the existing testing capacities can be used to test a significantly increased number of individuals. As a consequence, adequate tracing and quarantine strategies to reduce community transmission can be established and valuable epidemiological studies relying on accurate prevalence rates can be performed for the ongoing pandemic situation.

## 1 Introduction

The current spreading state of the COVID-19 pandemic urges authorities around the world to take measures in order to contain the disease or, at least, to reduce its propagation speed, as commonly referred to by the term “curve flattening”.^1^ At the time of writing, the World Health Organization (WHO) reported 3,917,366 cases and 274,361 deaths with 61,578 new cases in the last 24 hours.^2^ In particular, more than fifty countries experiencing larger outbreaks of local transmission and severe depletion of the workforce, for example, among healthcare workers (HCWs), had been reported to the WHO. Also, given the current number of tests described by several government agencies, this number likely underrepresents the total number of SARS-CoV-2 infections globally.

Even though a lot of research is currently being performed towards a cure of this infectious disease, to date, the most effective reasonable measure against its spread is the tracking and subsequent isolation of positive cases via an intensive test procedure on a large part of the population or at least important risk groups.^3^ A pilot study conducted by the University of Padua and the Italian Red Cross in Vò, Italy, showed encouraging results in this direction.^4^

At present, the standard tests for the detection of SARS-CoV-2, are nucleic acid amplification tests (NAAT), such as the quantitative reverse transcription polymerase chain reaction (qRT-PCR). These biochemical tests are based on samples from the lower respiratory or upper respiratory tract of tested individuals.^5^ The former is too delicate of an operation to be widely applicable and usually only feasible for hospitalized patients. In the routine laboratory diagnosis, however, sampling the upper respiratory tract with nasopharyngeal and oropharyngeal swabs is much less invasive and usually the method of choice.

The demand for this type of SARS-CoV-2 testing, however, is drastically increasing in many health care systems, resulting in shortages of necessary materials to conduct the test or capacity limits of the testing laboratories.^6^

As proposed by a large number of contributions (cf. Section 2 below), a promising way to make better use of the available capacities is to mix samples of different individuals before testing, and to first perform the test on these mixtures, the so-called *pools*, as if it were only one sample. When a pool tests negative, this is interpreted as a negative result for all pooled specimens. When a pool is tested positive, however, this only entails that it contains some infected specimen and further considerations are necessary.

This concept of *group testing* (also called pooled testing or pooling) goes back to mathematical ideas developed in the 1940’s and has since been used for tests based on various biospecimens such as swab, urine, and blood.^7-9^ In particular, group tests are employed when testing for sexually transmitted diseases such as HIV, chlamydia, and gonorrhea, see and references therein, and were recently used in viral epidemics such as influenza.^10,11^

Very recently, there have also been successful proofs of concept for experimental pooling strategies in SARS-CoV-2 testing. An Israeli research team demonstrated the feasibility of pooling up to 32 samples; they encountered false negative rates of around 10%.^12^ Subsequently, a German initiative filed a patent for a new approach that allows for so-called minipools combining 5-10 samples with a significantly reduced false negative rate.^13^ Similarly, a US American research group performed a test with 12 pools of 5 specimens, each from individuals at risk, and were able to correctly identify the two infected individuals out of the 60 with only 22 tests.^14^

The main goal of these works is to demonstrate the feasibility of the experimental design; they propose to use the original group testing design by Dorfman of including each specimen into exactly one pool then testing every specimen of the pool again individually in case of a positive outcome of the group test.^7^ Other works over the last weeks have suggested refined approaches, typically based on examples or, from a more theoretical viewpoint, with a simplified model.^15-21^

In this manuscript, we will demonstrate and systematically explore that even within the limitations of the initial experimental designs for COVID-19 testing, more sophisticated pooling strategies can lead to a significantly reduced number of tests. Thus connecting the recent SARS-CoV2 pool tests to the rich literature on group testing developed over the last decades may be a key ingredient for effectual national responses to the current pandemic.

Such connections have been established by Abdalhamid, Bilder and McCutchen by incorporating a decision step regarding how to optimize the number of samples within each pool based on the estimated infection rate – this led to the choice of 5 for the pool size.^14^ The problem of choosing the right pool size had previously been analyzed in many works.^22-24^

The theoretical and practical understanding of group testing developed since the first results of Dorfman,^7^ however, goes far beyond merely optimizing the pool sizes.^25,26^ For example, it is also possible to study group testing in the case of responses involving three categories or more,^27^ and to use pooling for the more involved problem of estimating the prevalence of a disease in a population.^22^

Two threads of development of particular relevance for SARS-CoV-2 testing concern *array testing* strategies in which every specimen is included in more than one pool and pooling strategies explicitly designed for joint testing of individuals of different risk groups, a special case of *informative testing*. We will explain these approaches in detail in Section 2 the main message of this paper is that in realistic prevalence regimes for the current COVID-19 pandemic, both of them may help to improve the testing efficiency even significantly beyond the gain achieved by the simple pooling strategies implemented in the first approaches. By no means we claim statistical originality; our goal is rather to explore and numerically compare classical methods for a variety of realistic parameter choices, demonstrating their efficiency for large scale SARS-CoV-2 testing. This paper is accompanied by a repository of source code that allows for parallel computation and comparative visualizations.^28^

The remainder of our paper is structured as follows. After discussing the fundamentals of group testing in Section 2, the constraints that should be taken into account in the model will be described in Section 3 as well as its practical limitations. Section 4 presents the numerical experiments and data visualizations and Section 5 concludes the report with a summary of our findings as well as a discussion of implications and directions for follow-up work.

## 2 Group Testing

As described in Section 1, group testing (GT) is the procedure of performing joint tests on mixtures of specimens, so-called pools, as a whole, instead of administering individual tests, thereby requiring significantly fewer tests than the number of specimens to be tested. Ideally, this joint test will produce a positive outcome if any one of the specimens in the pool is infected and a negative outcome otherwise. Because of the limited information contained in a positive outcome, it is required to test certain specimens multiple times - either in parallel for all the specimens, or sequentially with additional testing only for those specimens with positive test results.

Sequential test designs in which grouping of samples into pools in each stage depends on the results of the former stages, are called *adaptive*. For *non-adaptive* methods, in contrast, all the sample groupings are specified in advance, which translates into a one stage procedure in which all pool tests can be performed in parallel.

A special class of adaptive test designs are *hierarchical* tests, where in the first stage, each specimen is included in exactly one pool, and, in every subsequent stage, groups with positive results are divided into smaller non-overlapping groups and retested, while all specimens contained in groups with negative results are discarded. The original Dorfman test, for example, is a two-stage hierarchical group test.

The Figure 1 illustrates the hierarchical structure of the Dorfman test with a 10 x 10 illustrative microplate. Each circle in the plate represents specimens from separate individuals and the red circles are the infected ones that need to be identified. The specimens are then amalgamated row by row to perform a group test for each row. A positive test result indicates that some individual in the corresponding row is infected. Once the results from the group tests are available, they can be used for the next stage, so only the specimens sharing a pool with an infected specimen will need to be retested.

**Figure 1:**
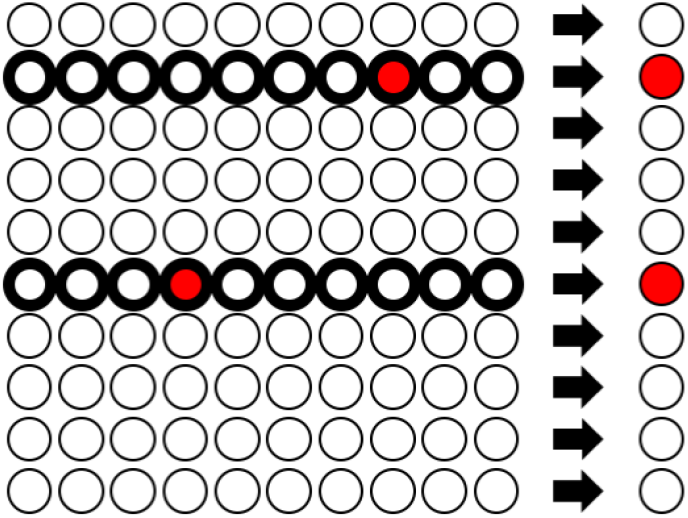
Hierarchical Testing as proposed by Dorfman. 100 specimens are randomly sorted in groups/rows of size 10. As indicated on the right-hand side, the row-wise group test correctly identifies the groups which contain a positive sample (indicated by the red color). Every sample of a positive group will be flagged as *possibly positive* (indicated by the bold circle) and used for the next stage of tests.

Entirely non-adaptive group testing procedures have been designed and analyzed using techniques at the interface of coding theory,^29^ information theory,^26^ and compressive sensing.^30-32^ The symbiosis among those fields lead to several development such as the establishment of optimal theoretical bounds for the best expected group testing strategies.^33^ However, some of the developments lead to algorithms that may not be practically efficient to implement and, consequently, are not suited for many medical applications including SARS-CoV-2 testing.

Nevertheless the idea of including every specimen in multiple pools to be tested in parallel is an integral part of many medical testing procedures, as the implementation of hierarchical tests with many stages can be rather complex and hard to automatize. Often, the test proceeds by arranging the specimens in a two-dimensional array and assembling all the specimens of each column in a pool, and then all the specimens of each row also in a pool.^34^ This testing strategy is a special instance of the so-called *array testing*, already mentioned in Section 1. In this way, every specimen is included in exactly two pools. All the specimens in the intersection of two pools with positive test results have to be retested in a second stage, but the number of these individual tests can be considerably smaller than for the Dorfman design. The Figure 2 illustrates the array testing procedure for a 10 × 10 microplate with two infected individuals; here only four of the 100 specimens need to be retested.

**Figure 2:**
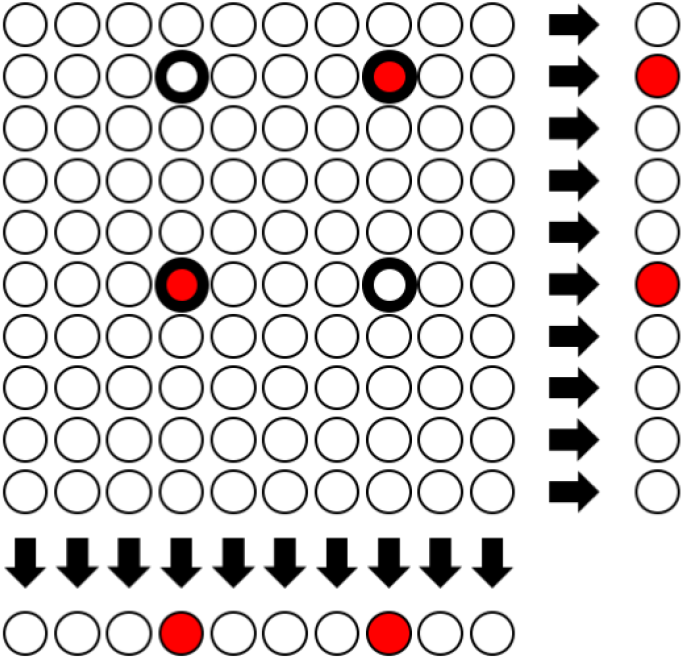
Array Testing. In addition to testing the row groups, column group tests are performed simultaneously. Only specimens which were tested positive in **both** group tests will be flagged as *possibly positive*. While this is an example with two simultaneous pool tests, also a higher amount of simultaneous tests can be performed.

Sometimes, for array tests, an initial *master pool* consisting of all specimens in a certain array is formed and all the *k*^2^ individuals are tested together. This allows for a rejection of a large group in case it exhibits a negative result. Otherwise one proceeds with the array strategy illustrated above. It is important to note, however, that master pooling should be used when there are no clear restrictions on the pool size, e.g., given by dilution effects. In case that such effects are not present, as claimed recently at least for small pool sizes,^13^ master pooling strategies could be explored.

Another important methodological advancement in group testing is the design of *informative tests*, i.e. testing strategies that are not based on the assumption of a uniform infection rate, but rather incorporate different estimates for the infection rate of subgroups of the population. We expect that such strategies will be of particular relevance for SARS-CoV-2 testing; for example the infection rate among health care professionals or elderly care workers is expected to be higher than for citizens working from home due to different levels of exposure, and similarly a stratification based on the level of symptoms also seems reasonable. A first attempt to make use of such a stratification for SARS-CoV-2 testing has recently been made with two subpopulations.^18^ This paper, however, only assembles homogeneous pools within the two subpopulations and hence does not make use of the full power of informative testing. Namely, the testing efficiency can be significantly improved by smartly assembling combined pools with members of both subpopulations.

Indeed our simulations confirm that this approach, when available, can help improve testing efficiency for realistic choices of parameters. At the same time, we expect that for best performance, one will have to employ a combination of different approaches.

As for many other applications, the design of the GT strategy needs to be driven by the following challenges.^35^

i. What practical considerations restrict the pooling strategies available to the laboratory?
ii. How do the pool size and the choice of assay for NAAT affect the ability of a pooling algorithm to detect infected individuals in a testing population?
iii. Given the assay and maximum pool size, what efficiencies can be expected for different pooling strategies in testing populations with different prevalences of the disease or well-defined subgroups of varying prevalence?
iv. How can pooling strategies be expected to impact the accuracy of the results?

Especially the fourth point has not received much attention in the literature on GT approaches for SARS-CoV-2 testing yet. Like most other testing procedures, qRT-PCR for COVID-19 misclassifies some negative specimens as positive and vice versa, as quantified by the *sensitivity* and the *specificity* of the test (the precise definitions are recalled in Section 3).

Causes of these inaccuracies that have been documented include *low viral load in some patients, difficulty to collect samples from COVID-19 patients, insufficient sample loading during qRT-PCR tests, and RNA degradation during sample handling process*.^36^ Some of these effects are to be amplified in group testing procedures, so it becomes even more important to take errors into account.

At the same time, the accuracy of a test is difficult to assess. Namely, as described above, NAAT is used to quantify the abundance of SARS-CoV-2 genetic material in a sample similarly to tests for other viral infections.^37^ In the specific case of qRT-PCR, the abundance measurement is on a continuous scale, the cycle (Ct) at which the readout, given by a fluorescence trace, surpasses a threshold. A decision boundary for a positive observation, i.e., infected, has to be established based on negative samples, i.e., biological control. Accordingly, the estimates on false negative and false positive rates of NAAT tests (and group tests in particular) for the SARS-CoV-2 infection depend on the strength of the classifier induced by this decision boundary. The accuracy of this classifier is influenced by a number of factors such as the following.

1. The ability of the test to selectively amplify virus genetic material depends on primer design. Multiple primers for qRT-PCR testing on COVID-19 samples were recently compared and found to be similarly strong, with a few exceptions of published weaker primers.^38^
2. A large worry about group testing is that the pooling of few positive samples with many negative samples could push the virus concentration in the pooled sample below the detection limit, increasing the false negative rate. This effect has been investigated by studying the test accuracy for dilutions containing virus samples, and false-negatives rates were found to be below 10% at a wide range of dilutions, suggesting that the qRT-PCR stage of the testing pipeline introduces small error rates only.^12^ Still, it is of fundamental importance to accurately estimate the errors introduced by dilution effects since a good understanding of the error is crucial to allow for any reliable inference in a disease study.^39^
3. Thirdly, sample extraction methods may have varying yield in virus material: This yield depends on the tissue or fluid that is sampled and on the processing of the sample, such as the time between sampling and qRT-PCR or the temperature at which the sample is held. One would expect this sample extraction to mostly have a destructive effect and to inflate negative rates rather than inflate positive rates.
4. The establishment of gold standard disease labels on samples that were also tested with NAAT is of fundamental importance to assess the overall accuracy of the classifier. There is little such data for COVID-19 testing right now. To this end, a recent study analyzed the positive test result rate of qRT-PCR tests on COVID-19 patients identified based on symptoms, where the symptom-based diagnosis served as a ground truth.^40^ They found false negative rates of individual tests of around 11% to 25% on sputum samples. At the same time, false-positive rates are hard to estimate in the current situation in which non-symptomatic infections occur at unknown frequency and because of the lack of reference gold-standard labels for positive observations that are non-symptomatic. However, as sample collection does likely contribute little to false positive rates, the overall false positive rate of a group test would largely depend on the qRT-PCR stage in which there is reason to believe that it should be small. Some previous studies on the use of PCR for similar infectious diseases such as SARS-like viruses as well as for SARS-CoV-2 reported high sensitivity for PCR.^38,41^ Indeed, in the absence of cell culture methods, qRT-PCR tests are considered to be the *gold standard* for the identification and diagnosis of most pathogen.

The importance of such estimates described above lead to a recent collaborative effort between FIND, a Swiss foundation for diagnostics, and the World Health Organization for the COVID-19 pandemic in order to evaluate the qRT-PCR tests and to assess their accuracy.^42^ FIND is currently evaluating a list of more than 300 SARS-CoV-2 tests commercially available and to establish accurate estimates for sensitivity and specificity with their respective confidence intervals.^43^ Based on the preliminary findings, in this work we will assume that the specificity of a single PCR test is 99%. For the sensitivity we will mostly assume the value of 99% as well but also explore the impact of lower values to account for potential dilution effects along the several tables presented in the appendix.

A common thread in the various aspects discussed in this section seems to be the large variety of relevant parameters due to differences between testing scenarios and uncertainty as a consequence of infected individuals without symptoms. In this note, we aim to illustrate that the test design of choice should very much depend on these parameters to make best use of the testing capacities. We will provide numerical comparison between different designs for large classes of parameters, such as the sensitivity, specificity, and the expected number of tests per person, so the design can be constantly adapted to what is the best fit to the currently best estimate of, e.g., the infection rate and the sensitivity.

Before discussing our numerical results, we will precisely introduce the relevant design parameters and testing strategies in the next Section.

## 3 Methods

### 3.1 Terminology

We start by introducing some terminology.

- **Prevalence** *p*: This is the assumed infection rate of the population which is going to be tested, that is, the fraction of the population that is infected. Hence it also is the probability of infection for a randomly selected individual. For simplicity of notation we will write *q* = 1 – *p* for the probability that a randomly selected individual is negative. When the test subjects can be divided into groups with different fractions of infected subjects, we also speak of the prevalences of these subgroups. Without further specification, however, the term refers to the full population to be tested.
- **Number of Stages:** This denotes how many steps the method performs sequentially and these steps are characterized by the fact that each stage requires the results from the previous one. In this paper, we will study adaptive methods with up to three stages, even though more stages, usually up to four in the case of infectious diseases, can be used.^34^
- **Divisibility:** This refers to the maximal number of tests that can be performed on a given specimen. This number provides a limitation on how many group tests can be performed, in parallel or in different stages, that include the corresponding test subject.
- **Group size** *k*: This is the size of the groups that are used in a pooling scheme. For a testing strategy to be feasible, one needs to ensure that the maximal group size *k* still allows for a reliable detection of a single positive in a pool of size *k*.
- **Sensitivity** *S_e_*: This is the probability that an individual test correctly returns a positive result when applied to a positive specimen or pool. A priori, this probability can be different depending on the number of positives included, for example due to dilution effects,^39,44,45^ but we will neglect this important distinction for the mathematical description below and assume that a PCR test has a fixed sensitivity independent of pool size. Analogously, for a pooling strategy *X*, *S_e_*(*X*) is the probability of the whole method *X* returning a positive result for a positive specimen.
- **Specificity** *S_p_:* This is the probability that an individual test correctly returns a negative result when applied to a negative specimen or pool. Again we assume that a PCR test has a fixed specificity independent of pool size. In case dilutions effects are taken into account and more specific information on how the sensitivity/specificity changes with the pool size *k* is added, one should write *S_e_* and *S_p_* with a dependency on *k*. Analogously, for a pooling strategy *X*, *S_p_*(*X*) is the probability of the method *X* returning a negative result when a negative specimen is tested.
- **Expected number of tests per person** *E*: We consider the *expected number of tests per person* as a measure of efficiency. Naturally, the expected number of tests per person of a method depends on the prevalence *p* as well as *S_e_* and *S_p_*, but also on the design parameters, such as the group size k and the number of stages. We will write *E*(*X*) to denote the expected number of tests per person for a method *X*, without explicitly indicating its dependence on these parameters for the sake of notational simplicity. There exist a recent discussions in the literature about alternative objective functions which take directly into account the effects on specificity and sensitivity. The findings, however, show that such alternative choice most often does not affect how the optimal group testing configuration.^46^

The optimal choice of design will depend on the aforementioned parameters. In Section 4, we will explore these dependencies numerically.

There is also some theory on the optimal design choice and the necessary amount of tests. An argument given by Sobel and Groll,^47^ which is based on the seminal works by Shannon and Huffman,^48,49^ shows the theoretical lower bound for the expected number of tests per individual of any given group testing method. More precisely, they showed that E (X) ≥ – *p* log *p* – *q* log *q* must hold for any method *X* with *S_e_*(*X*) = *S_p_*(*X*) = 1. In addition to its theoretical interest, it pragmatically indicates how much further improvement might still be possible. Note that it is only a bound, which may very well not be achievable with practically feasible methods. Figures 6 and 7 illustrate how the methods discussed here compare to this bound and how much gain one could expect for any large scale group testing strategy.

Regarding the influence of the infection rate, it has been established by Ungar that for infection rates 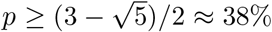, the optimal pool size is 1, so there *does not exist a group testing scheme* that is better than individual testing.^50^ Also, on a intuitive level, one may think that the higher the prevalence, the higher the expected number of tests should be. In fact, Yao and Hwang proved that the minimum of the expected number of tests with respect to all possible test strategies should be non-decreasing with respect to *p*, if 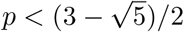.^51^

Therefore, in the COVID-19 pandemic where the prevalence in most countries, both among the tested individuals and the entire population is clearly believed to be smaller than the threshold provided by Ungar’s theorem, one can expect a significant reduction in average number of tests by employing suitable group testing methods. In the following subsection, we will discuss some of these methods and their mathematical formulation.

### 3.2 Standard group testing methods

In this subsection, we will recall some standard methods for group testing that we will numerically explore in the following section. An overview of these methods and their mathematical formulation can be found in the book by Kim and colleagues while their mathematical derivation were published by Johnson and colleagues.^52,53^

#### 3.2.1 2-stage hierarchical testing (D2)

Dorfman’s method is an adaptive method, which tests, in a first stage, each individual as part of a group of size *k*.^7^ Then, in the groups that tested positive, all the individuals are tested again individually in a second stage. Consequently, the test requires a divisibility of 2. The probability of a pool of size *k*, here denoted by ℙ*_k_*, drawn at random from the population to test positive is

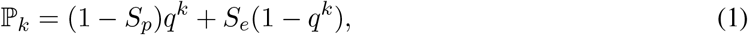

the expected number of tests per person of the method is given by

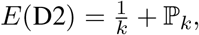

and its sensitivity and specificity are

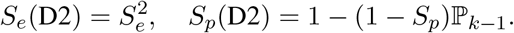

A slight improvement of Dorfman’s method is possible by omitting one of the individual tests per pool in the second stage and only performing it in a third stage when at least one of the other second-stage tests of that pool has a positive result - exploiting that if all test results in the second stage are negative, the last specimen must be infected for the group test to be valid.^47^

A more significant modification was proposed by Sterrett.^8^ In his method, the second stage is modified by performing individual tests until the first positive is found. Then a pooling procedure similar to the first stage is performed for the remaining, still unlabeled, specimens, and this scheme is repeated until all specimens are labeled. While requiring a smaller number of tests per individual on average, especially for small infection rates,^24^ the number of stages which need to be performed sequentially is not known a priori and may be very high. As such Sterrett’s method is more involved in practice, while D2 is a simple and straightforward procedure. Thus the latter is often preferred in applications, which is also why we will perform the simulations for the original form of D2 in this paper.

#### 3.2.2 3-stage hierarchical testing (D3)

In this method, each individual is tested as part of a pool of size *k* in a first stage. Every pool that tests positive is then split into subgroups, which are tested in a second stage. Every member of a subgroup with positive result in the second stage, is tested individually in a third stage. Consequently, this method requires divisibility 3. In this paper, we will focus on the case that all subgroups are of size *s*. Expected number of tests per person, sensitivity, and specificity of this method are given by

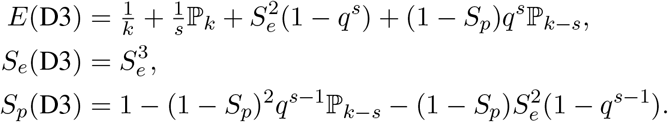

A schematic comparison between the hierarchical tests with two and three stages, D2 and D3, is given in Figure 3.

**Figure 3:**
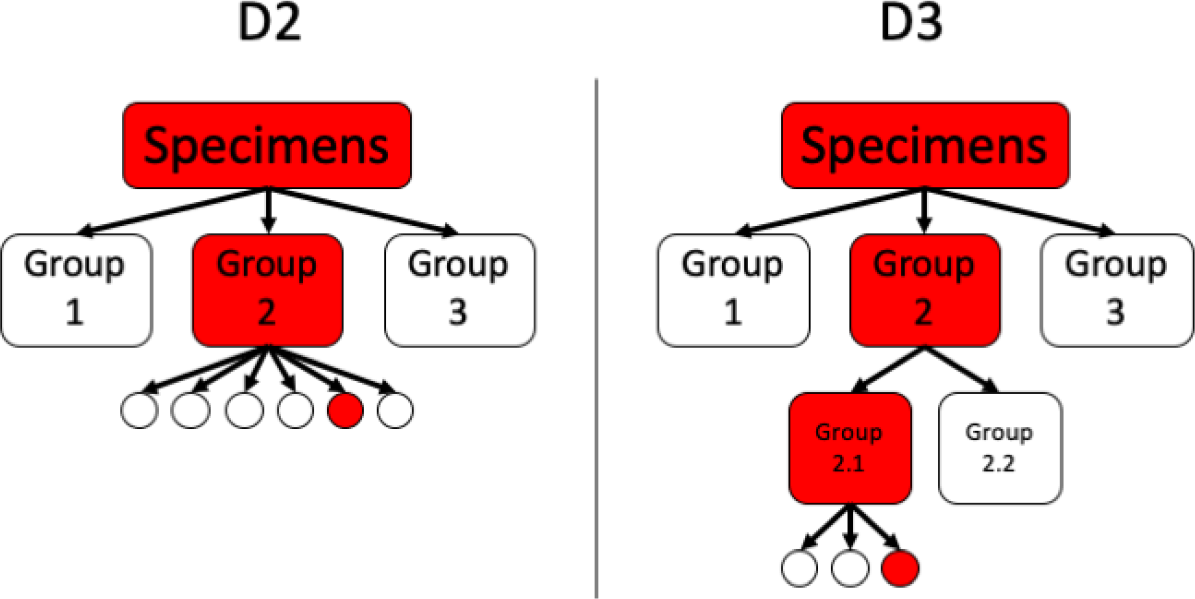
Comparison between D2 and D3

#### 3.2.3 Array testing (A2)

This is a 2-stage method, originally proposed by Phatarfod and Sudbury and later explored by Kim and colleagues,^52,54,55^ that tests every individual twice in a first stage as a part of two different groups of size *k*. In a second stage all the individuals, for which both group test results are positive, are tested individually. Consequently, this method requires divisibility 3.

Precisely determining the optimal way to assemble the pools is rather non-trivial, see, e.g., the publication by Kim and colleagues,^52^ but the following configuration provides a good trade-off between simplicity and expected number of tests. At first, *k*^2^ specimens are arranged in a *k* × *k* array, then every row and every column is pooled and subjected to a group test. This ensures that each specimen is tested exactly twice as part of a group of size *k* and constitutes the unique intersection of these two pools. For *S_p_* = *S_e_* = 1 it is sufficient to only test a person individually if both its row and column tests return positive results. In this case one obtains the following formula for the expected number of tests

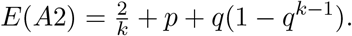

If *S_e_* or *S_p_* differ from 100%, the first stage may yield positive rows without any positive columns or vice versa. In this case it makes sense to test every member of such a row or column individually.^52,56^ This results in a slight increase in sensitivity at the expense of a slight increase in expected number of tests per person. As this change makes the formulas much more involved, we omit them here and refer to the corresponding literature.^52^

**Figure 4:**
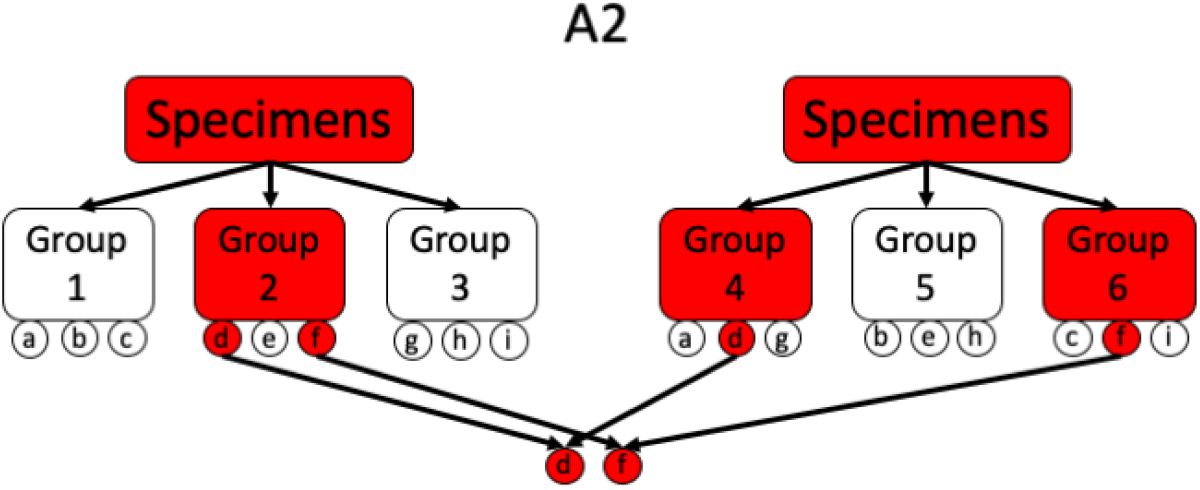
Illustration of a simple A2 procedure where the positive individuals are uniquely determinable after the first stage. Every individual *a,b,c, …,i* gets pooled exactly twice.

A2 can be generalized to procedures with three or more simultaneous pools. In this case, the pools could be assembled, for example, by creating pools along the diagonals and/or the anti-diagonals of an array, in addition to rows and columns.^56^ An advantage of such approaches is that the group tests for all these pools can be performed in parallel, which can lead to faster test results, but one has to take into account that the sensitivity is decreasing with the number of pool tests per individual.

The method above can be extended to higher-dimensional procedures, i.e., *j* > 3, and a connection to optimally efficient two-stage methods can be established. Note that these arrays have size *k^j^* rendering this approach practically infeasible very quickly as *j* and *k* increase. More concretely,^57^ showed that if the prevalence is *p* = 0·01, then an (almost) optimally efficient two-stage method can be achieved by *j* = 6 and *k* = 74, i.e., a 6-dimensional array with side length 74. However, the population, in this case, would need to contain 74^6^ ≈ 164 billion individuals to be screened and is impractical to be applied in any real world problem. Thus, the quest for methods that use the same principles but are effective for a realistic population size still remains.

**Figure 5:**
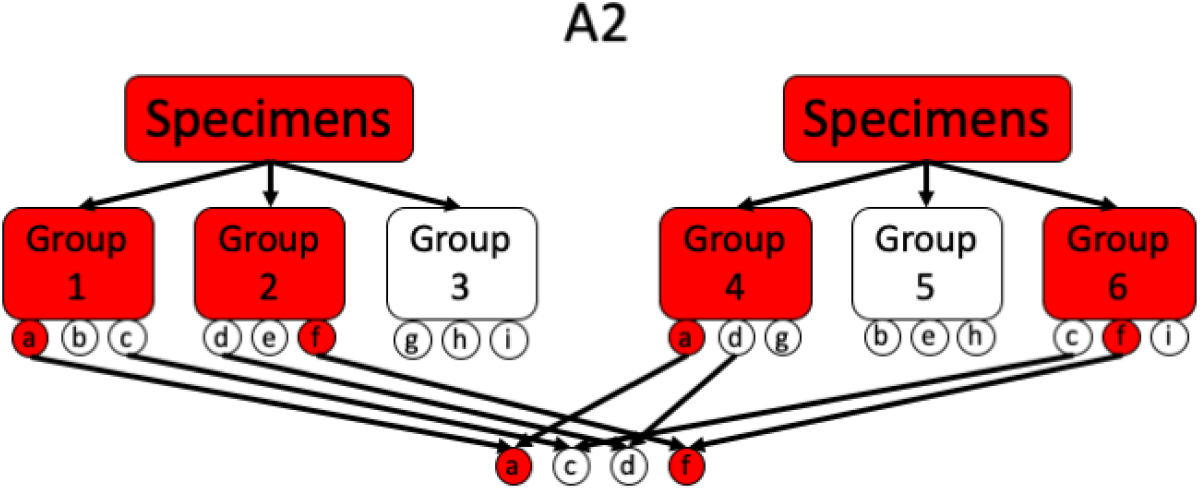
Illustration of a simple A2 procedure where also two negative samples got flagged the first stage.

#### 3.2.4 Non-adaptive array testing (A1)

All the group testing methods discussed so far terminate with an individual test for all specimens with positive test results in all previous stages to avoid false positives only based on the choice of the pools. In a situation with shortage of test components, there may be scenarios where one is willing to accept a significant number of additional false positives as a means to reduce the expected number of tests and simplify the test design - in particular, it is desirable to perform all different tests necessary for a testing procedure in parallel.

Towards this goal, one may consider replacing the last stage of individual tests in an adaptive procedure by an additional pooling dimension to be performed in parallel, hence transforming the adaptive into a non-adaptive method.

When this adaption is applied to the Dorfman method, one obtains a procedure A1_2_ that is identical to the first stage of A2. When applied to A2, this yields a method A1_3_ of three parallel pool tests per specimen, again without a decisive individual test at the end. By design, the resulting methods have a significantly lower specificity, but lead to a reduction in necessary tests. An additional advantage is that the resulting methods are fully non-adaptive and can be performed in a single testing stage, allowing for faster test results. At the same time, the adaptation from the methods D2 and A2 does not effect the divisibility required nor the sensitivity of the resulting procedure as adding another additional pooling dimension is accompanied by omitting the last stage - one is really just trading specificity for a lower number of tests and non-adaptivity.

Hence a suitable decision parameter is the minimal acceptable specificity. By the trade-off just mentioned, this also implicitly determines the group size and hence the expected number of tests per person via the relations

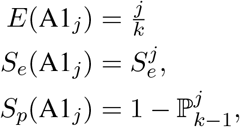

where *j* = 2,3. It is important to note that such tests can only be used when a certain false positive rate can be accepted. If a non-adaptive method with perfect detection of positive individuals, assuming perfectly accurate PCR, is required, a theoretical result by Aldridge shows that no test strategy is better than individual testing.^58^ Also, in contrast to the adaptive tests discussed above, the minimal number of expected tests per person alone is not a viable measure for the optimal choice of the group size *k* - it would yield a strong bias towards tests with many false positives. For the remainder of this work, the threshold for the minimal acceptable specificity is set to 95%. Nevertheless, we will give a short comparison with a preset of 90% and 97% in Section 4.

### 3.3 Extension to the informative case

As described in Section 2, it is possible to incorporate prior information such as demographic, clinical, spatial or temporal knowledge into refined estimates for the prevalence and to stratify the population accordingly, reflecting the heterogeneous distribution of the infected individuals. This heterogeneity, first explored by Nebenzahl and Sobel,^59^ and Hwang,^60^ can be exploited for refined GT strategies.

From a mathematical point of view, informative tests are somewhat more challenging to analyze.^61-64^ To illustrate the findings of the informative tests analysis and demonstrate its relevance for SARS-CoV-2 testing, we will work with a scenario where two distinct subpopulations, one with a high prevalence *p*_high_ (e.g. HCWs) and another, larger, subpopulation of individuals with low prevalence *p*_low_ (e.g., representative samples of the general population) are to be tested. As shown for example by Bilder and Tebbs,^65^ informative testing reduces the expected number of tests per individual even further when compared with their corresponding non-informative counterparts. As argued by them, it is crucial to exploit this heterogeneity and employ an efficient mixing strategy of individuals from both subpopulations to form the pools. Our goal here has a different perspective on how to exploit such strategies as will be discussed in the next section. It sheds light on testing methodologies where as much individuals as possible should be tested with the available tests while subject to the constraint of constantly testing high-risk individuals such as HCWs.

## 4 Numerical Results

In this section, we will numerically explore different design choices in group testing for SARS-CoV-2. A key tool is the R-package *binGroup* for identification and estimation using group testing, that features the computation of optimal parameter choices for standard group testing algorithms.^67^ We have complemented this package by a repository of source code for parallel computation and comparative visualization that has been used to create all the graphics in this section and is available for the reader to produce visualizations adapted to different prevalence ranges of interest.^28^

As indicated in the previous section, the choices of the correct method and the optimal group size *k* heavily depend on several constraints, most importantly the underlying prevalence *p* (or the subpopulation prevalences for a refined model). In this work, instead of attempting to find the optimal method we evaluate the properties of a group testing design for a single fixed group size. We will investigate different infection scenarios with the different group testing methods described above. We apply the tests D2, D3, A2, and A1*_j_* with overall prevalence varying from 0·25% to 15%. The results for D2, D3, and A2 have been simulated using *binGroup2* while A1*_j_* has been implemented separately.^66^

An important aspect to take into account when putting the amount of individuals tested per available test into perspective, is that methods based on multiple pools or stages will typically have a smaller overall sensitivity than individual tests, cf. Section 3.2. It is crucial to integrate the sensitivity considerations into any pooling strategy.^45^ In Tables 2, 3 and 4, we will illustrate (potential) efficiency increase assuming a sensitivity of 99% and 90%, respectively, for the qRT-PCR test. As mentioned before, extensive tests are currently being performed to confirm the high accuracy of qRT-PCR for SARS-CoV-2 testing. Indeed, they indicate that many available PCR procedures for SARS-CoV-2 testing show a sensitivity of or close to 100%.^43^ Nevertheless, an appropriate quantitative understanding of pooling effects and viral load progression on the sensitivity is still an active discussion.^68^

For a PCR sensitivity of 99%, we observe that the reduction caused by the use of a pooling method is very small (97% for D3, A2, and A1_3_; 98% for D2 and A1_2_). Only a single PCR procedure showed a low sensitivity of 90% when choosing a specific gene target (compared to 100% when choosing another target).^43^ In that case, we find a sensitivity of (73% for D3, A2, and A1_3_; 81% for D2 and A1_2_). While the specificity of PCR already appears to be close to 100%, the tables indicate that D2, D3, and A2 improve the specificity even further while A1_2_ and A1_3_ fulfill the preset threshold *S_p_*(.) ≥ 95%. Due to the specificity constraint, A1_2_ can not be recommended for very high infection rates of at least 12% as there is no reduction of necessary tests over individual testing. A1_3_ is more robust but shows the same behavior at *p* > 15%.

*S_e_*(.) and *S_p_*(.) depend mostly on the method and underlying sensitivity *S_e_* of the qRT-PCR test and barely change for increasing p. Therefore, Table 5 shows the change of *S_e_*(.) and *S_p_*(.) for *p* = 3% and varying S_e_. It should be noted that the sensitivity *S_e_*(.) virtually does not depend on the specificity *S_p_* of PCR. Only a slight change in initial group size can be detected. As explained in Section 3, the sensitivity can be computed as 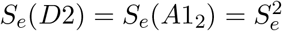 and 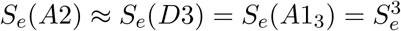.

To reflect practical considerations such as dilution effects,^12^ we constrain the group size to at most 16. We observe that all the methods yield a significantly reduced expected number of tests per person as compared to individual testing. This improvement decays with growing infection rate, in line with our discussion above. For prevalence values below 4%, and hence including the estimated range of current infection rates for SARS-CoV-2 in different countries,^69^ all adaptive methods (D2, D3, A2) allow to test at least 3 times as many individuals with the same amount of tests. Around a prevalence of 3% both non-adaptive methods allow to test around 5 individuals per test if a false positive rate up to 5% can be accepted.

Compared to individual testing where only a single individual can be tested per available test, Figures 6 and 7 demonstrate the average amount of individuals which can be tested per available test when applying different group testing methods. For infection rates as high as 2%, up to 5 times as many individuals compared to amount of available tests can be tested using adaptive methods. For a low prevalence below 0.5% this number varies between a 7 to 15-fold efficiency increase.

**Figure 6:**
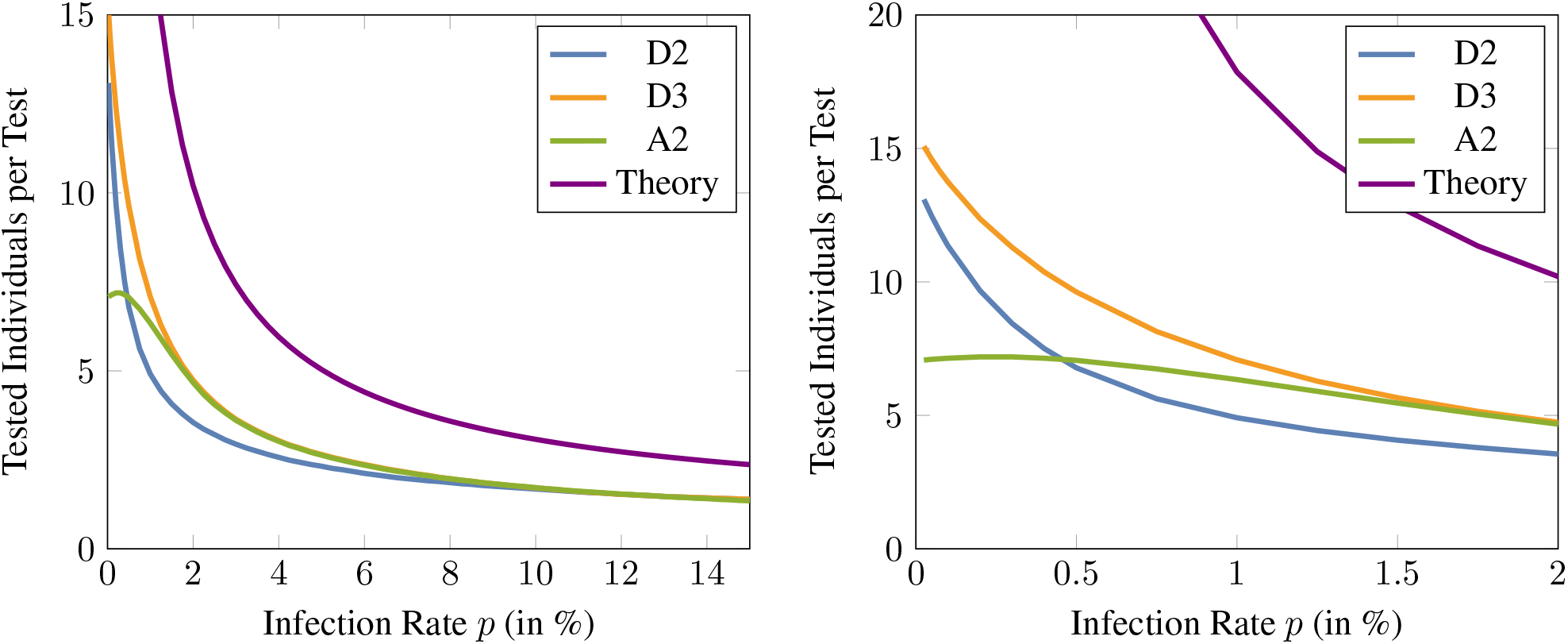
Number of individuals that can be tested per test available for the different adaptive methods. Here, the sensitivity and specificity are assumed to be 99%. The theoretical bound given by^47^ is also shown for a comparison and the maximum group size is assume to be 16. The figure on the right is a zoomed version of the left figure and illustrates the low prevalence regime of infection rates up to 2%.

**Figure 7:**
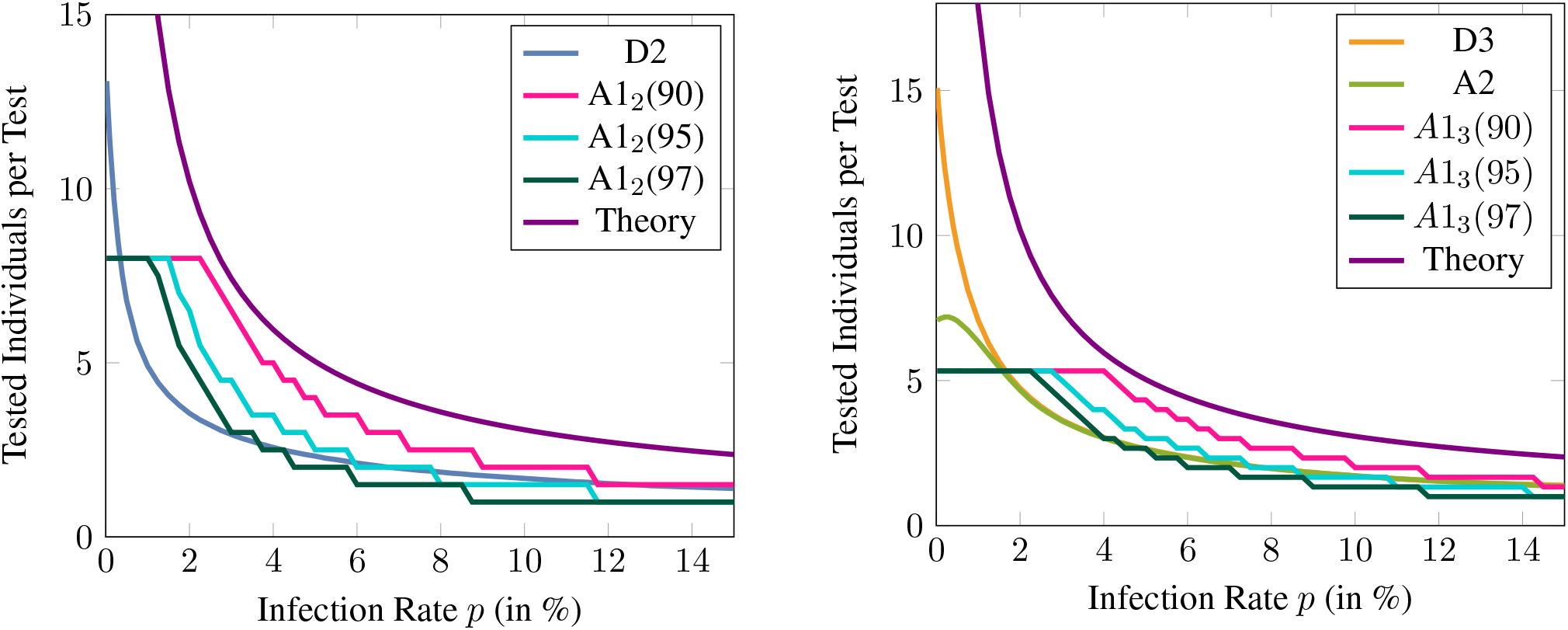
Number of individuals that can be tested per test available for different non-adaptive and their corresponding adaptive methods. A1*_j_*-9X denotes the non-adaptive method A1*_j_* with specificity threshold 9X%. Here, the sensitivity and specificity of qRT-PCR are assumed to be 99%. The theoretical bound given by^47^ is also shown for a comparison and the maximum group size is assume to be 16.

Figure 7 shows the efficiency improvement of A1*_j_* compared to the corresponding adaptive method. The specificity reduction, the biggest drawback of the proposed non-adaptive methods, is controlled by setting the threshold to 90%, 95% and 97%. Naturally, the methods relying on the lowest threshold show the biggest improvement. The suggested threshold of 95% leads to a significant improvement of A1_2_ compared to D2 for an infection rate between 0·4% and 5%. A1_3_ significantly exceeds A2 and D3 for a prevalence between 2·5% and 5%.

This is exemplified by some numerical examples in Table 1; for example, this entails that for an infection rate of 0·4%, the city of Munich with 147 million inhabitants could be tested with only 141 thousand tests using D3, the 6·69 million inhabitants of Rio de Janeiro could be tested using around 1 million tests if the infection rate does not exceed 10% and the adaptive methods D3 or A2 are performed. If a false positive rate up to 5% is considered acceptable, the non-adaptive method A1_2_ would only require 836,000 tests and at the same time allow for higher prevalence values of up to 1·5%.

**Table 1:** Illustration of the expected total amount of tests needed for testing in three different cities by assuming specificity and sensitivity of 99%. All numbers are in million·

| Munich (population: 1.47) |  |  |  |  |  |
| --- | --- | --- | --- | --- | --- |
| p | D2 | D3 | A2 | A1<sub>2</sub> | A1<sub>3</sub> |
| 0.025% | 0.112 | 0.097 | 0.207 | 0.184 | 0.276 |
| 0.05% | 0.118 | 0.101 | 0.207 | 0.184 | 0.276 |
| 0.075% | 0.123 | 0.104 | 0.206 | 0.184 | 0.276 |
| 0.1% | 0.129 | 0.107 | 0.206 | 0.184 | 0.276 |
| 0.2% | 0.151 | 0.119 | 0.204 | 0.184 | 0.276 |
| 0.3% | 0.173 | 0.131 | 0.204 | 0.184 | 0.276 |
| 0.4% | 0.196 | 0.141 | 0.206 | 0.184 | 0.276 |
| 0.5% | 0.218 | 0.153 | 0.209 | 0.184 | 0.276 |
| 1.0% | 0.298 | 0.207 | 0.232 | 0.184 | 0.276 |
| 1.5% | 0.362 | 0.260 | 0.269 | 0.184 | 0.276 |
| 2.0% | 0.413 | 0.310 | 0.315 | 0.226 | 0.276 |
| 2.5% | 0.459 | 0.357 | 0.363 | 0.294 | 0.276 |
| 3.0% | 0.500 | 0.404 | 0.407 | 0.326 | 0.294 |
| Vienna (population: 1.90) |  |  |  |  |  |
| p | D2 | D3 | A2 | A1<sub>2</sub> | A1<sub>3</sub> |
| 0.025% | 0.144 | 0.125 | 0.268 | 0.238 | 0.357 |
| 0.05% | 0.152 | 0.131 | 0.268 | 0.238 | 0.357 |
| 0.075% | 0.160 | 0.135 | 0.266 | 0.238 | 0.357 |
| 0.1% | 0.167 | 0.139 | 0.266 | 0.238 | 0.357 |
| 0.2% | 0.196 | 0.154 | 0.264 | 0.238 | 0.357 |
| 0.3% | 0.224 | 0.169 | 0.264 | 0.238 | 0.357 |
| 0.4% | 0.253 | 0.182 | 0.266 | 0.238 | 0.357 |
| 0.5% | 0.281 | 0.198 | 0.270 | 0.238 | 0.357 |
| 1.0% | 0.386 | 0.268 | 0.300 | 0.238 | 0.357 |
| 1.5% | 0.467 | 0.336 | 0.348 | 0.238 | 0.357 |
| 2.0% | 0.534 | 0.401 | 0.407 | 0.293 | 0.357 |
| 2.5% | 0.593 | 0.462 | 0.469 | 0.380 | 0.357 |
| 3.0% | 0.646 | 0.522 | 0.526 | 0.422 | 0.380 |
| Rio de Janeiro (population: 6.69) |  |  |  |  |  |
| p | D2 | D3 | A2 | A1<sub>2</sub> | A1<sub>3</sub> |
| 0.025% | 0.508 | 0.442 | 0.943 | 0.836 | 1.258 |
| 0.05% | 0.535 | 0.462 | 0.943 | 0.836 | 1.258 |
| 0.075% | 0.562 | 0.475 | 0.937 | 0.836 | 1.258 |
| 0.1% | 0.589 | 0.488 | 0.937 | 0.836 | 1.258 |
| 0.2% | 0.689 | 0.542 | 0.930 | 0.836 | 1.258 |
| 0.3% | 0.789 | 0.595 | 0.930 | 0.836 | 1.258 |
| 0.4% | 0.890 | 0.642 | 0.937 | 0.836 | 1.258 |
| 0.5% | 0.990 | 0.696 | 0.950 | 0.836 | 1.258 |
| 1.0% | 1.358 | 0.943 | 1.057 | 0.836 | 1.258 |
| 1.5% | 1.646 | 1.184 | 1.224 | 0.836 | 1.258 |
| 2.0% | 1.880 | 1.412 | 1.432 | 1.030 | 1.258 |
| 2.5% | 2.087 | 1.626 | 1.652 | 1.338 | 1.258 |
| 3.0% | 2.275 | 1.840 | 1.853 | 1.485 | 1.338 |

To summarize, below 1% infection rate, any of the presented group testing procedures will constitute an extreme improvement over individual testing while D3 shows the best performance. For 1% ≤ *p* < 6%, A2 and D3 show a comparable performance which is superior to D2. For *p* ≥ 10% all adaptive methods show a similar performance.

Considering the non-adaptive methods, A1_2_ requires a significantly reduced expected amount of tests for an infection rate between 1% and 4%. For a prevalence between 3% and 8%, *A*1_3_ shows the highest reduction in the number of tests of all methods. However, the trade-off between the lowest amount of tests and a false positive rate up to 5% has to be considered when choosing the testing method.

Next we numerically explore the average number of tests of different approaches for *informative testing*, with the goal of finding the best way to incorporate refined knowledge about different prevalences for distinct subpopulations. Each plot of Figure 8 compares the expected number of tests per person of two informative testing methods, namely the approach of choosing pools separately for the subpopulations, and the approach of assembling the pools with members of all subpopulations. We study a model with two subpopulations of different prevalence, and consider prevalence values between 5% and 25% for the high-risk and between 0·1% and 5% for the low-risk group. As far as we are aware, this assumption regarding different prevalence values for two groups, so in line with the two subpopulations we mention, was first mentioned in the context of SARS-CoV-2 by Deckert and colleagues,^18^ where they speak of homogeneous pools and use noninformative D2 for their analysis. However, the question of whether and how to adjust the testing procedure based on subpopulation knowledge did not arise in this work.

**Figure 8:**
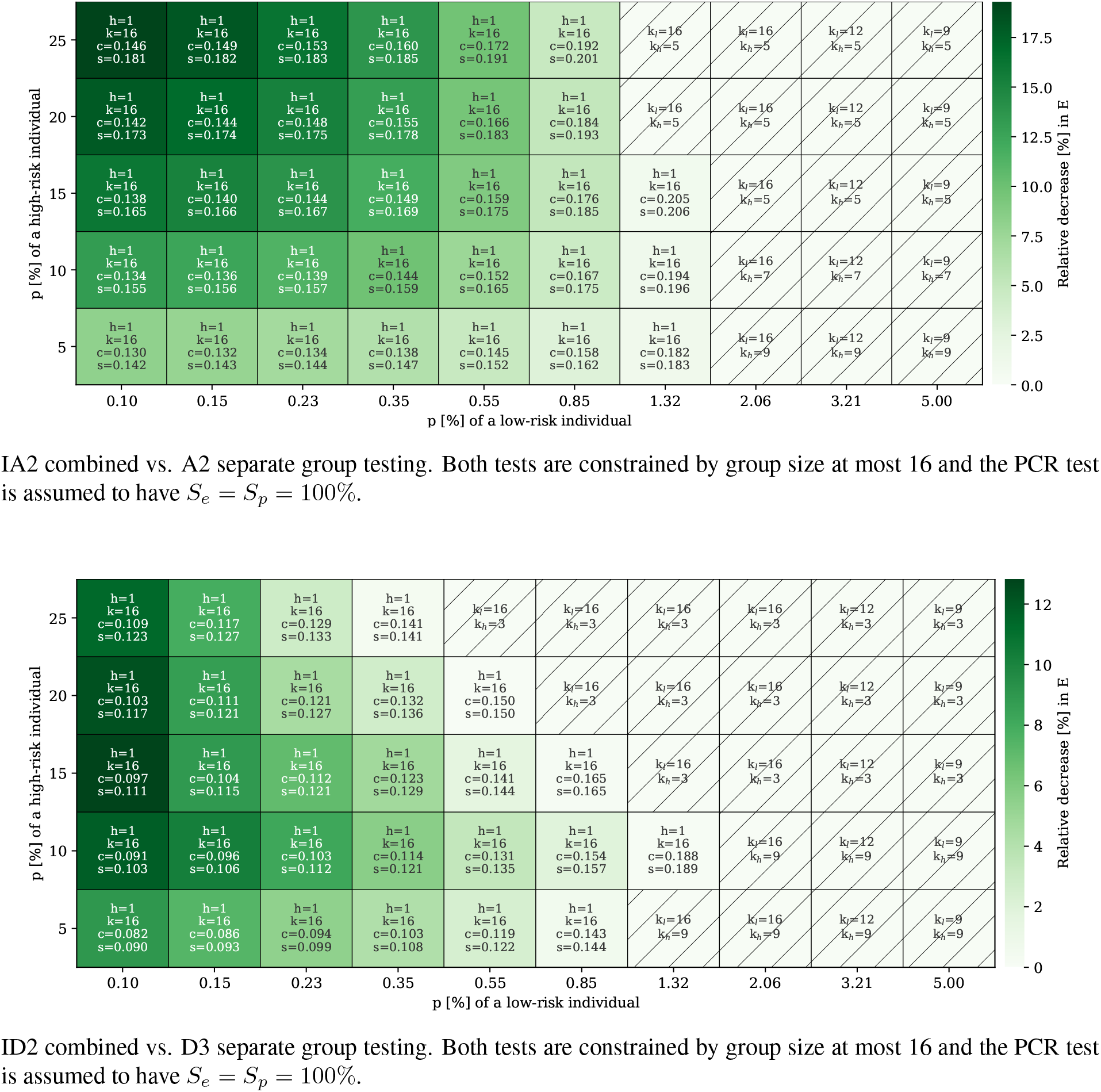
Comparison between different informative methods when the maximum group size is assumed to be 16. The dark green tiles indicate scenarios where pooling one individual of a high-risk group with specimens from low-risk groups reduces the average amount of tests needed over testing them separately. *h* represents the number of high-risk individuals per group, *c/s* is expected number of tests per person for combined/separate testing respectively and *k/k_l_/k_h_* are the optimal group sizes for combined/separate-low/separate-high testing. The hatched tiles indicate when this strategy should not be adopted.

We find that for A2 and D3, the advantage of assembling combined pools from both subpopulations gets larger when the prevalence of the low-risk group decreases. How it depends on the prevalence of the high-risk group differs depending on the methods and also the constraints imposed on the group size. For D2, however, the same phenomenon was not observed. More experiments of the same type but with different group sizes as well as different sensitivities and specificities can be visualized at our web application.^28^

## 5 Discussion

In this manuscript, we provide a comparison of general strategies for group testing in view of their application to medical diagnosis in the current COVID-19 pandemic.

Our numerical study confirms the recent observation that even under practical constraints for pooled SARS-CoV-2 tests, such as restrictions on the pool size, and for prevalence values in the estimated range of current infection rates in many regions,^69^ group testing is typically more efficient than individual testing and it allows for an efficiency increase of up to a factor 10 across realistic scenarios and testing strategies. We also find significant efficiency gaps between different group testing strategies in realistic scenarios for SARS-CoV-2 testing, highlighting the need for an informed decision of the pooling protocol. The repository for parallel computation and comparative visualization accompanying this manuscript allows the reader to visualize the performance of the different approaches similarly to the tables and graphics contained in this paper for different sets of parameters.^28^

For every scenario and method, an optimal pool size can be determined. However, the pool size is constrained biochemically by dilution effects and by sensitivity considerations. For a low prevalence, this can prevent choosing the optimal pool size. We find that within pooling protocols, sophisticated methods that employ multiple stages or multiple pools per sample, or exploit prevalence estimates for subpopulations have the strongest advantages at low prevalences.

Such low prevalence values are realistic assumptions especially for large scale tests of representative parts of the population, so these methods are particularly suited for full population screens or representative sub-population screens with the goal of reducing transmission and flattening the infection curve. This is of fundamental importance since transmission before the onset of symptoms has been commonly reported and asymptomatic cases seem to be very common.^70^ For example, 328 of the 634 positive cases on board of the formerly quarantined Diamond Princess cruise ship were asymptomatic at the time of testing, which corresponds to 52% of the cases. Another study conducted in a homeless shelter in Boston, MA, USA, confirmed that standard COVID-19 symptoms like cough, shortness of breath, and fever were uncommon among individuals who tested positive and strongly argues for universal PCR testing on that basis.^71^ Also, besides enhancing the tests of mild/asymptomatic cases, some disease control centers, such as the ECDC, recommend that group testing should potentially be applied to prevalence studies.^72^

The pooling schemes suggested here can also include routine tests of cohesive subpopulations with high prevalence, such as health care workers, and therefore propose a sensible way to include commonly available information about risk group into the setup.^73^ For certain scenarios, our numerical experiments show a reduced expected number of tests when employing combined pools consisting of high-risk and low-risk individuals provided some estimates for the prevalence in these two parts of the test population are available.

One could also envision separate pooled tests with different requirements on specificity and population coverage in sub-populations with different prevalence, again highlighting the importance of proper stratification: High specificity is for example likely desirable among health care workers whereas specificity may be partially traded for coverage during contact tracing. At the heart of these trade-offs lie considerations about the societal cost of false positives in comparison to the cost of missed diagnosis because of a lack of available tests.

The improved test efficiency of group testing is, however, only one aspect of test design. The practitioners face several issues when deciding if group testing can provide a feasible solution for massive tests procedures.^45^ Other important practical considerations are time constraints, specimen conservation for multi-stage testing, and resource availability, as well as the actual execution of the test in the labs, such as variation in pipetting and sample collection. All of them should be taken into account before the establishment of massive test policies.

qRT-PCR-based tests are currently widely deployed for COVID-19 diagnosis and, more generally, to identify current infections.^41,74^ As for any nucleic acid amplification tests, one can only identify cases where virus particles can still be detected. Thus for long-term disease monitoring, NAATs will have to be complemented by serological tests, as these can be used to infer the immunity state of a patient and hence identify past asymptomatic infections through detection of disease-specific antibodies. Such tests have already been deployed in a few cases.^75,76^ In contrast to the PCR testing procedures mainly discussed in this paper, the main intention of serological testing is to obtain accurate estimations of the number of unidentified previous infections as a measure for the progress towards herd immunity. Group testing can also be expected to yield accuracy gains for this problem. Namely, group testing for prevalence estimation is an active area of research with many recent advancements and we are confident that some of these results can be employed once pooled tests become available.^22,77^

In any case, there are still many well-established methodological tools available in the literature that have not yet been explored for SARS-CoV-2 testing, so we advocate for a continued exchange between theory, simulation and visualization, and practice.

## Data Availability

For comparative visualization and querying of the precomputed results we provide an interactive web application. The source code for computation is open and freely available.

https://gitlab.com/hararticles/group-testing-simulations

http://www.group-testing.com/

## Acknowledgement

The authors are grateful to Luciana Jesus da Costa, from the Virology Department at the Federal University of Rio de Janeiro, for insightful discussions about SARS-CoV-2.

## Contributors

PG and FK conceptualized the project. CMV, TF and FK conceptualized this paper; CMV and TF drafted it with fundamental contributions from PH, DE, DSF and JB. CMV, TF, DE and JB did the investigation for the connection between the virological application and the theoretical foundations of group testing. DSF gave important insights in the PCR testing processes. CMV and DE led the literature search while TF and CMV created the figures and tables used in the manuscript. PH, DE, and JB contributed with the idea, analysis and visualization of the combined testing and created the repository and the web application. PG, FJT and FK oversaw the study. All authors read and approved the final manuscript.

## Appendix

**Table 2:** Overview of the optimal group size (for each stage i) for different hierarchical methods and infection rates *p*. Besides *E*, the expected number of tests per person, *TIpT* gives the average amount of tested individuals per test. *S_e_*(·)/*S_p_*(·) denotes, respectively, the sensitivity/specificity of the method for optimal group size. (Assumption: *S_p_* = 0·99, **left** table: *S_e_* = 0·99, **right** table *S_e_* = 0·90)

| Method | $p$ | $k_1$ | $k_2$ | $k_3$ | $E$ | $TIpT$ | $S_e(\cdot)$ | $S_p(\cdot)$ | Method | $p$ | $k_1$ | $k_2$ | $k_3$ | $E$ | $TIpT$ | $S_e(\cdot)$ | $S_p(\cdot)$ |
| --- | --- | --- | --- | --- | --- | --- | --- | --- | --- | --- | --- | --- | --- | --- | --- | --- | --- |
| D2 | 0.025% | 16 | 1 | 0 | 0.076 | 13.09 | 98% | 100% | D2 | 0.025% | 16 | 1 | 0 | 0.076 | 13.14 | 81% | 100% |
|  | 0.05% | 16 | 1 | 0 | 0.08 | 12.45 | 98% | 100% |  | 0.05% | 16 | 1 | 0 | 0.08 | 12.56 | 81% | 100% |
|  | 0.075% | 16 | 1 | 0 | 0.084 | 11.88 | 98% | 100% |  | 0.075% | 16 | 1 | 0 | 0.083 | 12.03 | 81% | 100% |
|  | 0.1% | 16 | 1 | 0 | 0.088 | 11.35 | 98% | 100% |  | 0.1% | 16 | 1 | 0 | 0.087 | 11.55 | 81% | 100% |
|  | 0.2% | 16 | 1 | 0 | 0.103 | 9.67 | 98% | 100% |  | 0.2% | 16 | 1 | 0 | 0.101 | 9.94 | 81% | 100% |
|  | 0.3% | 16 | 1 | 0 | 0.118 | 8.44 | 98% | 100% |  | 0.3% | 16 | 1 | 0 | 0.114 | 8.75 | 81% | 100% |
|  | 0.4% | 16 | 1 | 0 | 0.133 | 7.50 | 98% | 99.9% |  | 0.4% | 16 | 1 | 0 | 0.128 | 7.82 | 81% | 99.9% |
|  | 0.5% | 15 | 1 | 0 | 0.148 | 6.78 | 98% | 99.9% |  | 0.5% | 16 | 1 | 0 | 0.141 | 7.09 | 81% | 99.9% |
|  | 1% | 11 | 1 | 0 | 0.204 | 4.91 | 98% | 99.9% |  | 1% | 11 | 1 | 0 | 0.194 | 5.15 | 81% | 99.9% |
|  | 1.5% | 9 | 1 | 0 | 0.246 | 4.07 | 98% | 99.9% |  | 1.5% | 9 | 1 | 0 | 0.234 | 4.27 | 81% | 99.9% |
|  | 2% | 8 | 1 | 0 | 0.281 | 3.55 | 98% | 99.9% |  | 2% | 8 | 1 | 0 | 0.268 | 3.73 | 81% | 99.9% |
|  | 2.5% | 7 | 1 | 0 | 0.312 | 3.21 | 98% | 99.9% |  | 2.5% | 7 | 1 | 0 | 0.297 | 3.36 | 81% | 99.9% |
|  | 3% | 6 | 1 | 0 | 0.34 | 2.94 | 98% | 99.9% |  | 3% | 7 | 1 | 0 | 0.324 | 3.09 | 81% | 99.8% |
|  | 4% | 6 | 1 | 0 | 0.37 | 2.70 | 81% | 99.8% |  | 4% | 6 | 1 | 0 | 0.37 | 2.70 | 81% | 99.8% |
|  | 5% | 5 | 1 | 0 | 0.411 | 2.43 | 81% | 99.8% |  | 5% | 5 | 1 | 0 | 0.411 | 2.43 | 81% | 99.8% |
|  | 6% | 5 | 1 | 0 | 0.447 | 2.24 | 81% | 99.8% |  | 6% | 5 | 1 | 0 | 0.447 | 2.24 | 81% | 99.8% |
|  | 7% | 5 | 1 | 0 | 0.481 | 2.08 | 81% | 99.8% |  | 7% | 5 | 1 | 0 | 0.481 | 2.08 | 81% | 99.8% |
|  | 8% | 4 | 1 | 0 | 0.512 | 1.95 | 81% | 99.8% |  | 8% | 4 | 1 | 0 | 0.512 | 1.95 | 81% | 99.8% |
|  | 9% | 4 | 1 | 0 | 0.54 | 1.85 | 81% | 99.8% |  | 9% | 4 | 1 | 0 | 0.54 | 1.85 | 81% | 99.8% |
|  | 10% | 4 | 1 | 0 | 0.566 | 1.77 | 81% | 99.8% |  | 10% | 4 | 1 | 0 | 0.566 | 1.77 | 81% | 99.8% |
|  | 11% | 4 | 1 | 0 | 0.592 | 1.69 | 81% | 99.7% |  | 11% | 4 | 1 | 0 | 0.592 | 1.69 | 81% | 99.7% |
|  | 12% | 4 | 1 | 0 | 0.616 | 1.62 | 81% | 99.7% |  | 12% | 4 | 1 | 0 | 0.616 | 1.62 | 81% | 99.7% |
|  | 13% | 4 | 1 | 0 | 0.64 | 1.56 | 81% | 99.7% |  | 13% | 4 | 1 | 0 | 0.64 | 1.56 | 81% | 99.7% |
|  | 14% | 4 | 1 | 0 | 0.663 | 1.51 | 81% | 99.7% |  | 14% | 4 | 1 | 0 | 0.663 | 1.51 | 81% | 99.7% |
|  | 15% | 4 | 1 | 0 | 0.685 | 1.46 | 81% | 99.7% |  | 15% | 4 | 1 | 0 | 0.685 | 1.46 | 81% | 99.7% |
| D3 | 0.025% | 16 | 8 | 1 | 0.066 | 15.08 | 97% | 100% | D3 | 0.025% | 16 | 8 | 1 | 0.066 | 15.17 | 72.9% | 100% |
|  | 0.05% | 16 | 6 | 1 | 0.069 | 14.58 | 97% | 100% |  | 0.05% | 16 | 8 | 1 | 0.068 | 14.71 | 72.9% | 100% |
|  | 0.075% | 16 | 6 | 1 | 0.071 | 14.14 | 97% | 100% |  | 0.075% | 16 | 6 | 1 | 0.07 | 14.33 | 72.9% | 100% |
|  | 0.1% | 16 | 6 | 1 | 0.073 | 13.74 | 97% | 100% |  | 0.1% | 16 | 6 | 1 | 0.072 | 13.97 | 72.9% | 100% |
|  | 0.2% | 16 | 4 | 1 | 0.081 | 12.36 | 97% | 100% |  | 0.2% | 16 | 6 | 1 | 0.079 | 12.72 | 72.9% | 100% |
|  | 0.3% | 16 | 4 | 1 | 0.089 | 11.29 | 97% | 100% |  | 0.3% | 16 | 4 | 1 | 0.086 | 11.70 | 72.9% | 100% |
|  | 0.4% | 16 | 4 | 1 | 0.096 | 10.37 | 97% | 100% |  | 0.4% | 16 | 4 | 1 | 0.092 | 10.85 | 72.9% | 100% |
|  | 0.5% | 16 | 4 | 1 | 0.104 | 9.62 | 97% | 100% |  | 0.5% | 16 | 4 | 1 | 0.099 | 10.12 | 72.9% | 100% |
|  | 1% | 16 | 4 | 1 | 0.141 | 7.08 | 97% | 100% |  | 1% | 16 | 4 | 1 | 0.131 | 7.63 | 72.9% | 100% |
|  | 1.5% | 16 | 4 | 1 | 0.177 | 5.66 | 97% | 100% |  | 1.5% | 16 | 4 | 1 | 0.162 | 6.18 | 72.9% | 100% |
|  | 2% | 16 | 4 | 1 | 0.211 | 4.74 | 97% | 99.9% |  | 2% | 16 | 4 | 1 | 0.191 | 5.23 | 72.9% | 100% |
|  | 2.5% | 16 | 4 | 1 | 0.243 | 4.11 | 97% | 99.9% |  | 2.5% | 16 | 4 | 1 | 0.219 | 4.56 | 72.9% | 99.9% |
|  | 3% | 12 | 3 | 1 | 0.274 | 3.64 | 97% | 99.9% |  | 3% | 16 | 4 | 1 | 0.246 | 4.06 | 72.9% | 99.9% |
|  | 4% | 12 | 3 | 1 | 0.329 | 3.04 | 97% | 99.9% |  | 4% | 12 | 4 | 1 | 0.296 | 3.38 | 72.9% | 99.9% |
|  | 5% | 9 | 3 | 1 | 0.377 | 2.65 | 97% | 99.9% |  | 5% | 12 | 4 | 1 | 0.341 | 2.93 | 72.9% | 99.9% |
|  | 6% | 9 | 3 | 1 | 0.423 | 2.37 | 97% | 99.9% |  | 6% | 9 | 3 | 1 | 0.381 | 2.63 | 72.9% | 99.9% |
|  | 7% | 9 | 3 | 1 | 0.466 | 2.15 | 97% | 99.9% |  | 7% | 9 | 3 | 1 | 0.418 | 2.39 | 72.9% | 99.9% |
|  | 8% | 9 | 3 | 1 | 0.507 | 1.97 | 97% | 99.9% |  | 8% | 9 | 3 | 1 | 0.453 | 2.21 | 72.9% | 99.9% |
|  | 9% | 9 | 3 | 1 | 0.546 | 1.83 | 97% | 99.8% |  | 9% | 9 | 3 | 1 | 0.487 | 2.05 | 72.9% | 99.9% |
|  | 10% | 9 | 3 | 1 | 0.584 | 1.71 | 97% | 99.8% |  | 10% | 9 | 3 | 1 | 0.519 | 1.93 | 72.9% | 99.8% |
|  | 11% | 9 | 3 | 1 | 0.619 | 1.61 | 97% | 99.8% |  | 11% | 9 | 3 | 1 | 0.549 | 1.82 | 72.9% | 99.8% |
|  | 12% | 4 | 1 | 1 | 0.652 | 1.53 | 98% | 99.7% |  | 12% | 9 | 3 | 1 | 0.578 | 1.73 | 72.9% | 99.8% |
|  | 13% | 3 | 1 | 1 | 0.678 | 1.47 | 98% | 99.8% |  | 13% | 9 | 3 | 1 | 0.606 | 1.65 | 72.9% | 99.8% |
|  | 14% | 3 | 1 | 1 | 0.7 | 1.43 | 98% | 99.7% |  | 14% | 9 | 3 | 1 | 0.633 | 1.58 | 72.9% | 99.8% |
|  | 15% | 3 | 1 | 1 | 0.722 | 1.39 | 98 % | 99.7% |  | 15% | 12 | 3 | 1 | 0.658 | 1.52 | 72.9% | 99.8% |

**Table 3:** Overview of the optimal group size (for each stage *i*) for array testing and infection rates *p*. Besides *E*, the expected number of tests per person, *TIpT* gives the average amount of tested individuals per test. *S_e_*(·)/*S_p_*(·) denotes, respectively, the sensitivity/specificity of the method for optimal group size (Assumption: *S_p_* = 0·99, **left** table: *S_e_* = 0·99, **right** table *S_e_* = 0·90)

| Method | $p$ | $k_1$ | $k_2$ | $k_3$ | $E$ | $TIpT$ | $S_e(\cdot)$ | $S_p(\cdot)$ | Method | $p$ | $k_1$ | $k_2$ | $k_3$ | $E$ | $TIpT$ | $S_e(\cdot)$ | $S_p(\cdot)$ |
| --- | --- | --- | --- | --- | --- | --- | --- | --- | --- | --- | --- | --- | --- | --- | --- | --- | --- |
| A2 | 0.025% | 16 | 1 | 0 | 0.142 | 7.07 | 98.6% | 100% | A2 | 0.025% | 16 | 1 | 0 | 0.142 | 7.04 | 86.1% | 100% |
|  | 0.05% | 16 | 1 | 0 | 0.141 | 7.10 | 98.5% | 100% |  | 0.05% | 16 | 1 | 0 | 0.142 | 7.04 | 85.4% | 100% |
|  | 0.075% | 16 | 1 | 0 | 0.14 | 7.12 | 98.4% | 100% |  | 0.075% | 16 | 1 | 0 | 0.142 | 7.05 | 84.8% | 100% |
|  | 0.1% | 16 | 1 | 0 | 0.14 | 7.14 | 98.4% | 100% |  | 0.1% | 16 | 1 | 0 | 0.142 | 7.05 | 84.1% | 100% |
|  | 0.2% | 16 | 1 | 0 | 0.139 | 7.19 | 98.1% | 100% |  | 0.2% | 16 | 1 | 0 | 0.142 | 7.05 | 82% | 100% |
|  | 0.3% | 16 | 1 | 0 | 0.139 | 7.19 | 97.9% | 100% |  | 0.3% | 16 | 1 | 0 | 0.142 | 7.03 | 80.2% | 100% |
|  | 0.4% | 16 | 1 | 0 | 0.14 | 7.14 | 97.7% | 100% |  | 0.4% | 16 | 1 | 0 | 0.143 | 6.99 | 78.8% | 100% |
|  | 0.5% | 16 | 1 | 0 | 0.142 | 7.06 | 97.5% | 100% |  | 0.5% | 16 | 1 | 0 | 0.144 | 6.94 | 77.6% | 100% |
|  | 1% | 16 | 1 | 0 | 0.158 | 6.34 | 97.2% | 100% |  | 1% | 16 | 1 | 0 | 0.156 | 6.43 | 74.5% | 100% |
|  | 1.5% | 16 | 1 | 0 | 0.183 | 5.46 | 97.1% | 100% |  | 1.5% | 16 | 1 | 0 | 0.175 | 5.72 | 73.5% | 100% |
|  | 2% | 16 | 1 | 0 | 0.214 | 4.67 | 97% | 99.9% |  | 2% | 16 | 1 | 0 | 0.2 | 5.01 | 73.1% | 99.9% |
|  | 2.5% | 14 | 1 | 0 | 0.247 | 4.05 | 97% | 99.9% |  | 2.5% | 16 | 1 | 0 | 0.228 | 4.38 | 73% | 99.9% |
|  | 3% | 13 | 1 | 0 | 0.277 | 3.61 | 97% | 99.9% |  | 3% | 14 | 1 | 0 | 0.256 | 3.91 | 73% | 99.9% |
|  | 4% | 11 | 1 | 0 | 0.331 | 3.02 | 97% | 99.9% |  | 4% | 12 | 1 | 0 | 0.306 | 3.27 | 73% | 99.9% |
|  | 5% | 10 | 1 | 0 | 0.381 | 2.63 | 97% | 99.9% |  | 5% | 11 | 1 | 0 | 0.351 | 2.85 | 73% | 99.9% |
|  | 6% | 9 | 1 | 0 | 0.426 | 2.35 | 97% | 99.9% |  | 6% | 10 | 1 | 0 | 0.392 | 2.55 | 73% | 99.9% |
|  | 7% | 8 | 1 | 0 | 0.468 | 2.14 | 97.1% | 99.8% |  | 7% | 9 | 1 | 0 | 0.43 | 2.32 | 73.1% | 99.8% |
|  | 8% | 7 | 1 | 0 | 0.509 | 1.97 | 97.1% | 99.8% |  | 8% | 8 | 1 | 0 | 0.467 | 2.14 | 73.2% | 99.8% |
|  | 9% | 7 | 1 | 0 | 0.545 | 1.83 | 97.1% | 99.8% |  | 9% | 8 | 1 | 0 | 0.5 | 2.00 | 73.1% | 99.8% |
|  | 10% | 7 | 1 | 0 | 0.582 | 1.72 | 97% | 99.8% |  | 10% | 7 | 1 | 0 | 0.534 | 1.87 | 73.2% | 99.8% |
|  | 11% | 6 | 1 | 0 | 0.616 | 1.62 | 97.1% | 99.8% |  | 11% | 7 | 1 | 0 | 0.563 | 1.78 | 73.1% | 99.8% |
|  | 12% | 6 | 1 | 0 | 0.648 | 1.54 | 97.1% | 99.8% |  | 12% | 7 | 1 | 0 | 0.593 | 1.69 | 73.1% | 99.8% |
|  | 13% | 6 | 1 | 0 | 0.68 | 1.47 | 97.1% | 99.8% |  | 13% | 7 | 1 | 0 | 0.622 | 1.61 | 73% | 99.7% |
|  | 14% | 6 | 1 | 0 | 0.712 | 1.41 | 97% | 99.7% |  | 14% | 6 | 1 | 0 | 0.649 | 1.54 | 73.2% | 99.8% |
|  | 15% | 5 | 1 | 0 | 0.742 | 1.35 | 97.1% | 99.8% |  | 15% | 6 | 1 | 0 | 0.674 | 1.48 | 73.2% | 99.7% |

**Table 4:**
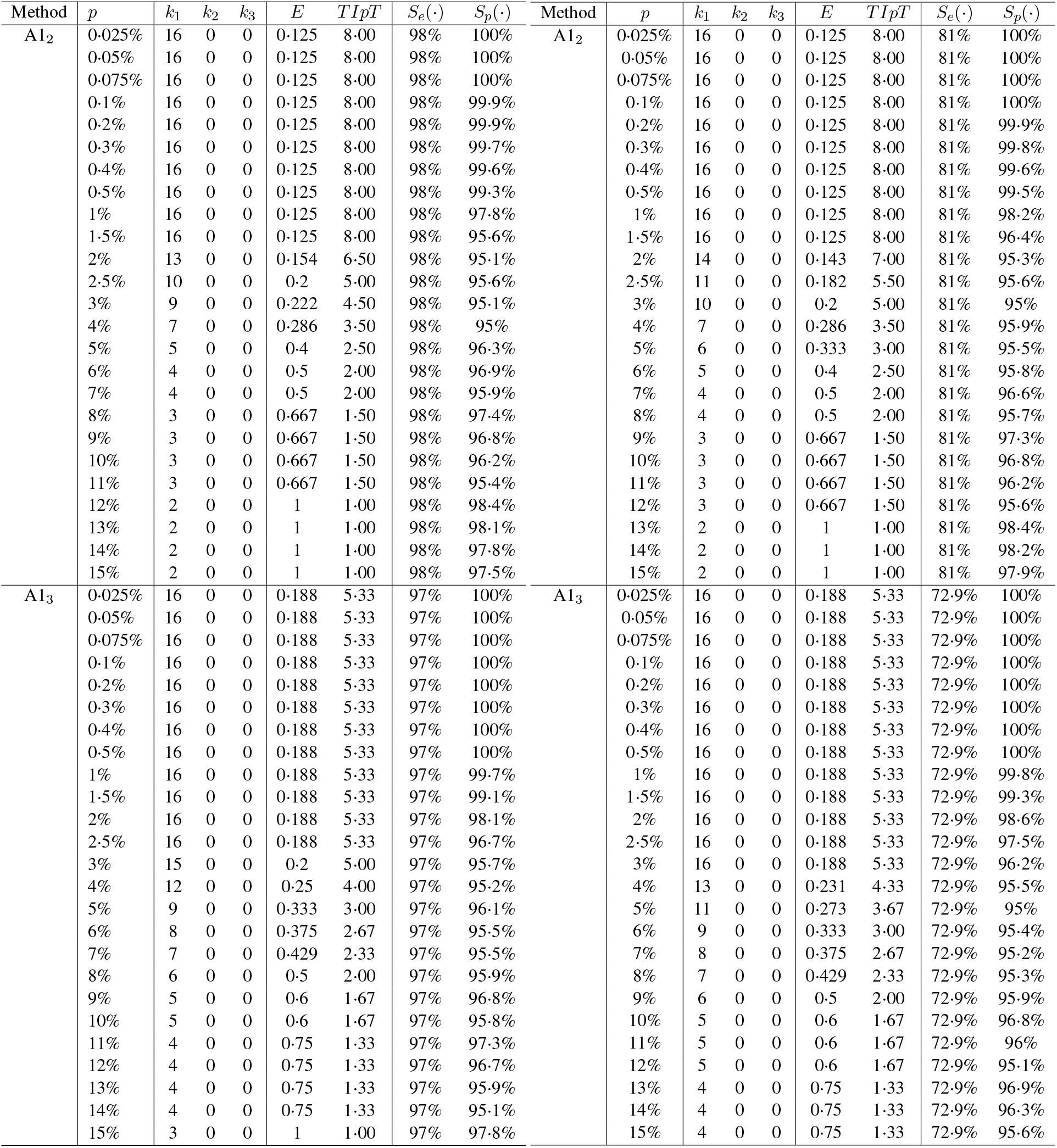
Overview of the optimal group size (for each stage i) for different non-adaptive methods and infection rates *p*. Besides *E*, the expected number of tests per person, *TIpT* gives the average amount of tested individuals per test. *S_e_*(·)/*S_p_*(·) denotes, respectively, the sensitivity/specificity of the method for optimal group size. (Assumption: *S_p_* = 0·99, **left** table: *S_e_* = 0·99, **right** table *S_e_* = 0·90)

**Table 5:** Overview of the sensitivity *S_e_*(·) and specificity *S_p_*(·) for different *S_e_* of qRT-PCR with a fixed prevalence assumption of *p* = 3% and specificity *S_p_* = 0·99%.

| Method | $S_e$ | $S_e(\cdot)$ | $S_p(\cdot)$ | $k_1$ |
| --- | --- | --- | --- | --- |
| D2 | 100% | 100% | 99.9% | 6 |
|  | 99% | 98% | 99.9% | 6 |
|  | 98% | 96% | 99.9% | 6 |
|  | 97% | 94.1% | 99.9% | 6 |
|  | 96% | 92.2% | 99.8% | 7 |
|  | 95% | 90.2% | 99.8% | 7 |
|  | 94% | 88.4% | 99.8% | 7 |
|  | 93% | 86.5% | 99.8% | 7 |
|  | 92% | 84.6% | 99.8% | 7 |
|  | 91% | 82.8% | 99.8% | 7 |
|  | 90% | 81% | 99.8% | 7 |
| D3 | 100% | 100% | 99.9% | 12 |
|  | 99% | 97% | 99.9% | 12 |
|  | 98% | 94.1% | 99.9% | 16 |
|  | 97% | 91.3% | 99.9% | 16 |
|  | 96% | 88.5% | 99.9% | 16 |
|  | 95% | 85.7% | 99.9% | 16 |
|  | 94% | 83.1% | 99.9% | 16 |
|  | 93% | 80.4% | 99.9% | 16 |
|  | 92% | 77.9% | 99.9% | 16 |
|  | 91% | 75.4% | 99.9% | 16 |
|  | 90% | 72.9% | 99.9% | 16 |
| A2 | 100% | 100% | 99.9% | 13 |
|  | 99% | 97% | 99.9% | 13 |
|  | 98% | 94.2% | 99.9% | 13 |
|  | 97% | 91.3% | 99.9% | 13 |
|  | 96% | 88.5% | 99.9% | 13 |
|  | 95% | 85.8% | 99.9% | 13 |
|  | 94% | 83.2% | 99.9% | 13 |
|  | 93% | 80.5% | 99.9% | 14 |
|  | 92% | 77.9% | 99.9% | 14 |
|  | 91% | 75.5% | 99.9% | 14 |
|  | 90% | 73% | 99.9% | 14 |
| A1<sub>2</sub> | 100% | 100% | 96% | 8 |
|  | 99% | 98% | 95.1% | 9 |
|  | 98% | 96% | 95.2% | 9 |
|  | 97% | 94.1% | 95.3% | 9 |
|  | 96% | 92.2% | 95.4% | 9 |
|  | 95% | 90.2% | 95.5% | 9 |
|  | 94% | 88.4% | 95.5% | 9 |
|  | 93% | 86.5% | 95.6% | 9 |
|  | 92% | 84.6% | 95.7% | 9 |
|  | 91% | 82.8% | 95.8% | 9 |
|  | 90% | 81% | 95% | 10 |
| A1<sub>3</sub> | 100% | 100% | 95.6% | 15 |
|  | 99% | 97% | 95.7% | 15 |
|  | 98% | 94.1% | 95.1% | 16 |
|  | 97% | 91.3% | 95.3% | 16 |
|  | 96% | 88.5% | 95.4% | 16 |
|  | 95% | 85.7% | 95.5% | 16 |
|  | 94% | 83.1% | 95.7% | 16 |
|  | 93% | 80.4% | 95.8% | 16 |
|  | 92% | 77.9% | 95.9% | 16 |
|  | 91% | 75.4% | 96.1% | 16 |
|  | 90% | 72.9% | 96.2% | 16 |

## Footnotes

By combining different diagonals resp. anti-diagonals in a suitable way, such that one gets groups of size k obeying the unique intersection property.

While working on this manuscript, the updated package *binGroup2* with improved and unified functionalities was released.^66^ Even though some of our calculations were performed with it, since the repository makes use of the previous version, we kept it here for consistency.^28^

One could expect that the expected number of tests per person of a group testing method does not improve for an increasing prevalence. Nevertheless, Table 3 indicates the opposite for low infection rates and sensitivity *S_e_* = 0.99. While for 0.1% prevalence 0.140 tests per individual are expected, this improves to *E* = 0.139 for *p* = 0.2%. The explanation for such oscillatory pattern comes from the general implementation of A2’s expected number of tests per person in the binGroup package^67^ where plausibility checks are done. As mentioned in Subsection 3.2.3 for *S_e_, S_p_ <* 1, in a scenario where a positive row/column group but not a single positive column/row group are found, individual tests of the positive row/column group should be performed. In a low prevalence setting, those additional tests have a higher impact and lead to the oscillation. As a side note, from a theoretical perspective, the oscillations in the expected number of tests per person for low prevalence do not contradict the theorem by Yao and Hwang^51^ since the theorem concerns the minimum over all possible strategies and this one, even though can be a very good method for the current purposes, does not achieve the theoretical minimum.

